# PERCEIVED STIGMATIZATION, SOCIODEMOGRAPHIC FACTORS, AND MENTAL HEALTH HELP-SEEKING BEHAVIORS AMONG PSYCHIATRIC OUTPATIENTS ATTENDING A PSYCHIATRIC HOSPITAL IN LAGOS, NIGERIA

**DOI:** 10.1101/2020.11.10.20229120

**Authors:** Ifeanyichukwu Anthony Ogueji, Taiye Emmanuel Ojo, Taiwo Nurudeen Gidado

## Abstract

Among the general population of patients with mental illness is a sub-population (psychiatric outpatients) who often encounter limited mental health help-seeking behaviors due to many unknown factors. Therefore, this study aimed to explore some predictors of mental health help-seeking behaviors among psychiatric outpatients. This cross-sectional study accidentally recruited 42 psychiatric outpatients receiving treatment at the Federal Neuropsychiatric Hospital, Yaba, Lagos State, Nigeria. Their mean age was 27.03±7.05 years (age range = 18-48 years). Data were collected using standardized questionnaires, and analyzed using SPSS (v. 22). Statistical significance set at *p<*.05. The first finding showed a positive but not significant relationship between perceived stigmatization and mental health help-seeking behavior. Second showed that gender had no significant influence on mental health help-seeking behavior. Third showed that age had a positive but not significant relationship with mental health help-seeking behavior. Last finding submitted that clinical diagnosis, religious affiliation, marital status, and educational qualification had a significant joint prediction on mental health help-seeking behaviour, with 28% variance explained. Only religious affiliation had a significant independent prediction. Our findings have practical implications for enhancing mental health help-seeking behavior and strengthening an interdisciplinary approach to mental health care.

## INTRODUCTION

Psychiatric outpatients are a sub-population in the general population of patients with psychiatric illness. This sub-population is often limited from seeking mental health help, unlike their inpatient counterparts who are less limited because of the treatment they receive within the health facility setting (Chaudhry, et al. 2017; Coker, et al 2019). The extent of mental health help-seeking behavior of this sub-population remains among the crucial factors explaining their mental health (Kodama, et al. 2016; Magaard, et al. 2017; Khiari, et al. 2019; Shi, et al. 2020). Also, mental health help-seeking behavior is linked with psychiatric comorbidities in this sub-population (Magaard, et al. 2017; Dongxin, et al. 2019). These, therefore, makes researching into the predictors of mental health help-seeking behavior among the psychiatric outpatients worthy of attention from mental health researchers.

The mental health help-seeking behaviors of psychiatric outpatients remain restricted despite that related studies have found factors such as willingness to seek professional help for a serious illness, feeling comfortable talking about personal problems with professionals, encouragement, and pressure to seek help by family or friends, mental health literacy, self-concept, and employment status, as factors predicting mental health help-seeking behaviors among psychiatric outpatients (Mojtabai, et al. 2016; Bitman-Heinrichs, 2017; Wigand, et al. 2019; Egwuonwu, et al. 2019). These submissions from related studies, therefore, creates a paucity of information on the predictive roles of perceived stigmatization and sociodemographic factors on mental health help-seeking behaviors among the population of interest.

Therefore, the current study aimed to investigate the predictive impacts of perceived stigmatization, and sociodemographic factors on mental health help-seeking behaviors among psychiatric outpatients. Perceived stigmatization and sociodemographic factors are very worthy of research attention and this is given the fact that these variables are recurrent in the lived experiences of psychiatric outpatients (Makanjuola, et al. 2016; Knack, et al. 2017; Heydari, et al. 2019). However, there are very limited data to validate the impacts of these variables on the help-seeking behaviors of our population of interest.

For instance, some studies have found that if faced with mental health troubles, people may conceal the mental health problems within themselves rather than seek professional help, to avoid anticipated stigmatization or rejection from society (Adewuya & Makanjuola, 2008; D’Avanzo, et al., 2012; Smolak, et al., 2013; Sheikh, et al. 2015; Pietkiewicz & Bachryj, 2016; Mantovani, et al. 2017; Ogueji & Constantine-Simms 2019; Ogueji & Okoloba, 2020). In another finding, sociodemographic factors such as educational qualification, gender, and age were associated with help-seeking for mental illness (Mackenzie, et al. 2006; Adewuya & Makanjuola 2008; Oladipo & Blogun 2010; Cornally & McCarthy, 2011; Mishra, et al., 2011; Wendt & Shafer, 2015; Kumuyi, 2017; Olarenwaju, et al. 2019). However, most of these findings were often limited in their applicability to psychiatric outpatients. Additionally, most of these studies suggested the need for extending their research to other sub-populations of psychiatric patients such as psychiatric outpatients.

### Theoretical Underpinning and Application

#### Threats-to-Self-Esteem Model

Fisher, et al. (1983) argued that mental health help-seeking is influenced by the extent to which the help-seekers perceive the help situation as a threat to their ego. Thus, if the help-seeker perceives the help situation as a threat to their ego, they are less likely to seek help. For instance, if people consider that they will be stigmatized when they seek help, they are likely not to attempt help-seeking or they are likely to try another route of help-seeking that does not bring stigmatization. Similarly, if people consider social factors that are present during help-seeking to be a threat to their ego, they are likely not to attempt help-seeking or they are likely to try another route of help-seeking. In essence, help-seeking according to the threats-to-self-esteem model, is determined by the extent to which the help-seeker considers the help a threat to his/her ego.

#### Research Questions

Given the overall aim of this study, and the threats-to-self-esteem model which is the theoretical framework for this study, the following research questions were thus asked:

1. What is the influence of perceived stigmatization on mental health help-seeking behavior among psychiatric outpatients?
2. What is the influence of gender on mental health help-seeking behavior among psychiatric outpatients?
3. What is the influence of age on mental health help-seeking behavior among psychiatric outpatients?
4. What are the contributions of clinical diagnosis, religious affiliation, marital status, and educational qualification to mental health help-seeking behavior among psychiatric outpatients?

#### Hypotheses

The following hypotheses were tested in this research:

1. There will be a significant relationship between perceived stigmatization and mental health help-seeking behavior among psychiatric outpatients.
2. There will be a significant gender difference in mental health help-seeking behavior among psychiatric outpatients.
3. There will be a significant association between age and mental health help-seeking behavior among psychiatric outpatients.
4. Clinical diagnosis, religious affiliation, marital status, and educational qualification will be significant joint and independent predictors of mental health help-seeking behavior among psychiatric outpatients.

## METHOD

### Design

A cross-sectional survey design was used to obtain data from the participants. The independent variables were perceived stigmatization and sociodemographic factors, whereas the dependent variable was mental health help-seeking behavior. To be enrolled in this study, willing participants were required to be adults (18 years or greater), an outpatient registered as receiving treatment at the Federal Neuropsychiatric Hospital, Yaba, Lagos State, Nigeria. It was also required that willing participants could communicate (written & verbal) using the English language because our instruments were available in English Language only. We also required that willing participants were in full or partial contact with reality. The third author (a clinical psychologist) was responsible for screening participants on the inclusion criteria.

### Participants and Setting

Accidental sampling technique was employed to recruit 42 participants in Federal Neuropsychiatric Hospital, Yaba, Lagos State, Nigeria. The hospital was established in 1907 and is among the oldest psychiatric facility in Nigeria. The high number of psychiatric outpatients with diverse demographics that are being cared for at the hospital, justified our choice for selecting the Federal Neuropsychiatric Hospital, Yaba, Lagos State, Nigeria as our study setting.

The mean age of participants was 27.03±7.05 years (age range = 18-48 years). Of the total participants, 31 were males, whereas 11 were females. Their distribution by religious affiliation showed that 26 were Christians, 14 identified with Islam, 1 identified with other religious affiliation, and 1 reported none. The marital status distribution showed that 32 were single, whereas 10 were married. The distribution by highest educational qualification showed that 31 had attained education from tertiary institutions, 10 had a high school education, and 1 had primary school education. The clinical diagnosis of participants showed that 22 were diagnosed with substance abuse induced schizophrenia, 6 were diagnosed with depression, 4 were diagnosed with anxiety disorders, 3 were diagnosed with simple schizophrenia, 3 were diagnosed with bipolar affective disorder, 2 were diagnosed with mania, 1 was diagnosed with dependent personality disorder, and 1 was diagnosed with paranoid schizophrenia.

### Measures

The standardized questionnaire utilized in this study comprised: demographic data of the participants, and questions relating to perceived stigmatization and mental health help-seeking behavior. Demographic variables included gender, age, marital status, religious affiliation, highest education attained, and clinical diagnosis of the patient. The stigma scale for mental illness was used to measure the perceived stigmatization and this was developed by King, et al. (2007). This scale was purposely chosen for the fact that the authors have demonstrated the relevance of the scale in understanding the role of the stigma of psychiatric illness in clinical research. It consists of 28 items. Illustrative items on the scale were; “Very often I feel alone because of my mental health problems”, “I am scared of how other people will react if they find out about my mental health problems”, or “People have been understanding of my mental health problems”. The scale is rated on a 5-point Likert format, i.e., 0= strongly disagree, 1= disagree, 2= neither agree nor disagree, 3= agree, 4= strongly agree. The internal consistency of the stigma scale as reported by the scale authors was 0.87. We found an internal consistency of 0.85 as determined by Cronbach’s alpha.

The mental health help□seeking behavior scale, adopted from Egwuonwu, et al. (2019), consists of 10 items, and this was used to assess the mental health help-seeking behaviors of participants. Illustrative items on the scale were; “I would want to get psychiatric attention if I was worried or upset for a long period of time”, “A person with an emotional problem is not likely to solve it alone, he/she is likely to solve it with professional help”, or “The idea of talking about problems with a psychologist is a good way to get rid of emotional conflicts”. Items were rated using the bipolar format (Yes or No). The minimum possible score was 0 while the maximum possible score was 10. A score of 50% or more was interpreted as good mental health help-seeking behavior whereas less than 50% was interpreted as poor mental health help-seeking behavior. Internal consistency of 0.78 was found in our study using Cronbach’s alpha.

### Procedure

The first author prepared a letter of ethical approval request and proposal that was submitted by the third author to the Research and Ethics Committee of the hospital to obtain ethical approval. After approval was granted, data collection from participants was proceeded to, based on the willingness and availability of participants. Before data collection, all participants were briefed about the research purpose, they were assured of the confidentiality of their responses, and only those that consented were given a consent form to confirm their willingness to be involved in the study.

The third author was responsible for data collection, and the administration of questionnaires occurred almost weekly (between late December 2019 and February 2020). At the end of each week, questionnaires were retrieved from participants, and all participants were thanked for participating in the study. Out of 300 questionnaires that were printed (based on consultation with 2 principal psychologists working at the study setting), 50 questionnaires were administered and retrieved; however, 42 questionnaires were completed. The “limitations of study” section of this paper has addressed why 50 questionnaires could be administered. As a part of the ethical consideration, the hospital management was informed that the outcome of the study would be communicated to them for policy action purposes. Finally, the IBM SPSS statistics (v. 22.0) was used for data analysis, and statistical significance was set at *p*<.05.

## RESULTS

### Hypothesis 1

First hypothesis was stated that there will be a significant relationship between perceived stigmatization and mental health help-seeking behavior among participants. This was tested using Pearson r.

**Table I.** Summary table of Pearson r showing the relationship between perceived stigmatization and mental health help-seeking behavior.

|  | 1 | 2 |
| --- | --- | --- |
| 1 PS | — |  |
| 2MHB | 0.12 | — |
| Mean | 93.71 | 7.83 |
| SD | 16.23 | 2.07 |

**PS = Perceived stigmatization**
**MHB = Mental health help-seeking behavior**

The above result showed that there was a positive relationship between perceived stigmatization and mental health help-seeking behavior at (*r* = 0.12; *df* = 40; *p* > 0.05). However, the relationship was not significant. Therefore, the stated hypothesis that there will be significant relationship between perceived stigmatization and mental health help-seeking behavior is falsified in this study.

### Hypothesis 2

It was stated that there will be a significant gender difference in mental health help-seeking behavior among participants. This was tested using independent sample t-test.

**Table II.** Summary of independent sample t-test table showing the gender differences in mental health help-seeking behavior.

| DV | Group | N | Mean | SD | T | Df | P |
| --- | --- | --- | --- | --- | --- | --- | --- |
|  | Male | 31 | 7.97 | 2.12 |  |  |  |
| Mental health help-seeking behavior |  |  |  |  | 7.28 | 40 | >.05 |
|  | Female | 11 | 7.45 | 1.97 |  |  |  |

From the above table, it is shown that male participants scored a higher mean score when compared with female counterparts on mental health help-seeking behavior. However, the difference was not significant as determined by independent sample t-test at [*t* (40) = 7.28; *p*>.05]. Consequently, the result falsifies the stated that there will be a significant gender difference in mental health help-seeking behavior among participants.

### Hypothesis 3

Third hypothesis stated that there will be a significant association between age and mental health help-seeking behavior among participants. This was tested below, using the Pearson r.

**Table III.**
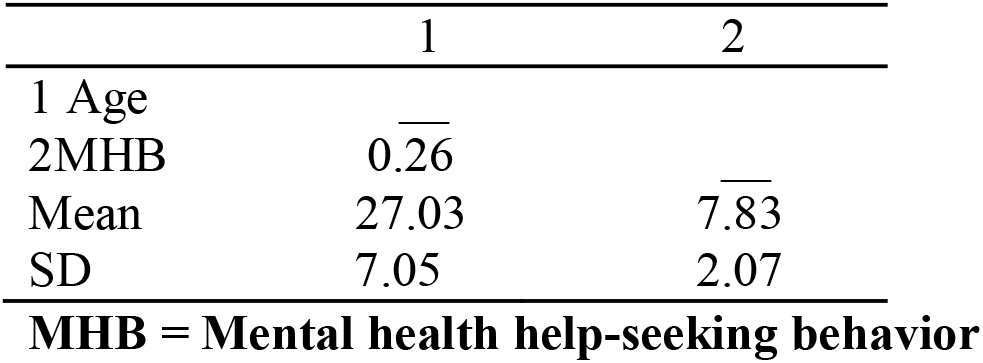
Summary table of Pearson r showing the relationship between age and mental health help-seeking behavior.

|  | 1 | 2 |
| --- | --- | --- |
| 1 Age | — |  |
| 2MHB | 0.26 | — |
| Mean | 27.03 | 7.83 |
| SD | 7.05 | 2.07 |

**MHB = Mental health help-seeking behavior**

The above result showed that there was a positive relationship between participants’ age and mental health help-seeking behavior at (*r* = 0.26; *df* = 40; *p* > 0.05). However, the relationship was not significant. Therefore, the stated hypothesis that there will be significant relationship between age and mental health help-seeking behavior is falsified in this study.

### Hypothesis 4

It was hypothesized that, clinical diagnosis, religious affiliation, marital status and educational qualification will be significant joint and independent predictors of mental health help-seeking behavior among participants.

**Table IV.** Summary of Multiple Regression Table, Showing the Independent and Joint Predictors of Mental Health Help-Seeking Behaviours.

| Predictors | B | T | P | R | R<sup>2</sup> | F | P |
| --- | --- | --- | --- | --- | --- | --- | --- |
| Clinical diagnosis | .21 | 1.46 | >.05 |  |  |  |  |
| Religious affiliation | -.37 | -2.47 | <.05 |  |  |  |  |
| Marital status | -.19 | -1.24 | >.05 |  |  |  |  |
|  |  |  |  | .53 | .28 | 3.57 | <.05 |
| Educational qualification | .07 | 0.46 | >.05 |  |  |  |  |

Criterion variable = Mental health help-seeking behaviour

The above table showed that clinical diagnosis, religious affiliation, marital status and educational qualification had a significant joint prediction on mental health help-seeking behaviour [*R* = .53; *R*^*2*^ = .28; *F* (4, 37) = 3.57; *p* < .05]. Furthermore, it was observed that the predictor variables jointly accounted for 28% variance in mental health help-seeking behavior (*R* =.53; *R*^*2*^ = .28), whereas other variables not considered in this study accounted for the remaining 72% variance.

The independent contribution of each predictor variable showed that of all the four (4) predictors, religious affiliation had a significant independent contribution on mental health help-seeking behavior at (*β* = -.37, *t* = -2.47; *p* < .05), whereas other predictors (clinical diagnosis, marital status, and educational qualification) did not have significant independent contributions. Given these, the stated hypothesis that; clinical diagnosis, religious affiliation, marital status and educational qualification will be significant joint and independent predictors of mental health help-seeking behavior among participants is partly accepted in this study.

## DISCUSSION AND IMPLICATION

We conducted a cross-sectional study to determine some predictors of mental health help-seeking behaviors among psychiatric outpatients attending a psychiatric facility in Lagos, Nigeria. Our first finding showed that there was no significant relationship between perceived stigmatization and mental health help-seeking behavior. Second, we found that there was no significant gender difference in mental health help-seeking behavior. Third, we found that there was no significant relationship between age and mental health help-seeking behavior. Fourth, we found that clinical diagnosis, religious affiliation, marital status, and educational qualification had a significant joint prediction on mental health help-seeking behavior, with a 28% variance in mental health help-seeking behavior explained by the predictors. In the same fourth finding, it was observed that of all the predictor variables, religious affiliation had a significant independent prediction on mental health help-seeking behavior.

The implication of the above findings suggests that the biopsychosocial context (such as clinical diagnosis, religion, marital status, or educational qualification) of psychiatric outpatients play a very key role in explaining their mental health help-seeking behaviors. Such roles are that the biopsychosocial context of psychiatric outpatients could strengthen or weaken their mental health help-seeking behaviors. Therefore, the biopsychosocial context of the study population is worthy of attention during mental health care. The implication of our findings agrees with related studies that submitted that the psychological and sociocultural context of living in Black societies (e.g., Nigeria) where our current study was conducted, can influence peoples’ health-enhancing behaviors concerning their mental health (Ogueji & Okoloba, 2020).

Further, our first finding disagreed with related studies where perceived stigmatization was found to influence peoples’ intention to seek mental health help (Sheikh, et al. 2015; Ogueji & Constantine-Simms, 2019). Our first finding also disagreed with the threats-to-self-esteem model that guided our study (Fisher, et al. 1983). Additionally, the first finding contradicted the submissions from a sample of participants in England which showed that stigmatization influenced help-seeking for mental illness (Mantovani, et al. 2017). It also contradicted the findings of a qualitative study in Rome that revealed that stigmatization influenced help-seeking behaviors (Pietkiewicz & Bachryj, 2016).

Because our first finding showed a positive (although not significant) relationship between perceived stigmatization and mental health help-seeking behavior; it, therefore, raised a hypothesis as to whether participants were motivated to seek professional help despite that they were stigmatized because they found solace against mental illness stigma in the psycho-education that they received during therapy or in the compassionate characteristics of their therapist(s). Future research can benefit from this hypothesis.

The second finding in our study disagreed with Olarenwaju, et al. (2019) who submitted that gender was a significant predictor of mental health help-seeking behavior. Our second finding also disagreed with the findings from an American survey which submitted that males were more likely than females to engage in health-enhancing behaviors concerning mental health (Wendt & Shafer, 2015). On the other hand, the second finding agreed with Kumuyi (2017) who argued that gender was not a significant predictor of mental health help-seeking behavior; and the consensus could be explained by the fact that the current study and Kumuyi’s (2017) study were conducted on a similar sample (i.e., psychiatric outpatients). Additionally, our second finding aligned with a study conducted with 710 participants in Italy (D’Avanzo, et al., 2012). The study found no significant gender difference in mental health help-seeking behavior.

Further, the third finding in our study disagreed with Oladipo and Blogun (2010) who found that age significantly predicted the mental health help-seeking behaviors of people. The third finding also disagreed with Mackenzie, et al. (2006) who found that age was significantly associated with help-seeking for mental illness. On the other hand, our fourth finding was consistent with a related study where variables similar to clinical diagnosis, religious affiliation, marital status, and educational qualification were found to significantly predict mental health help-seeking behaviors (Adewuya & Makanjuola 2008). Our fourth finding also supported a study by Ward et al. (2013) that found that the religion of African Americans impacted their mental health help-seeking behaviors. Our fourth finding also supported the study of other scholars in America, Smolak, et al. (2013) that reported religion to play an important role in mental health help-seeking behavior. Additionally, a study conducted with 72 adults in an Irish community reported a significant contribution of educational qualification to help-seeking behavior (Cornally & McCarthy, 2011). A study conducted in North India supported our fourth finding by reporting the impact that religious beliefs had on mental health help-seeking behavior (Mishra, et al., 2011). Further, a qualitative study with Catholic clergy in Rome supported our fourth finding by reporting that religion impacted the help-seeking behavior of participants (Pietkiewicz & Bachryj, 2016). The agreement between our findings and the findings from other cultures (e.g., western worlds), may suggest that our study has practical benefits for contributing to help-seeking for mental illness among the study population in other cultures (however, future cross-cultural studies can benefit from this argument). On the other hand, the disagreement between our findings and the findings from other cultures may suggest the impact of cultural differences, or the need for further studies that examine the reason for the disagreement.

From our findings, we recommend that intervention programs that are aimed at strengthening the mental health help-seeking behaviors of psychiatric outpatients should be cautious of the significant predictors identified in our study. Giving attention to the significant predictors reported in our study is expected to strengthen an interdisciplinary approach to mental health care because of the biological, psychological, and sociocultural predictors of mental health help-seeking behaviors that were found in our study. Finally, our study is an extension of related studies because related studies often found the predictors of mental health help-seeking behaviors from a sociocultural perspective (e.g., Mackenzie, et al. 2006; Wendt & Shafer, 2015; Kumuyi, 2017; Mantovani, et al. 2017; Ogueji & Constantine-Simms, 2019; Ogueji & Okoloba, 2020).

## LIMITATIONS OF THE STUDY

Our study could not access a large sample size because most psychiatric outpatients in the study setting were not in stable mental health conditions to complete the questionnaires when we conducted this study. Therefore, future studies can draw a large sample size from multiple psychiatric facilities simultaneously. Another factor that limited our access to large sample size was the timing of our study (i.e., our study was conducted when the threats of COVID-19 pandemic newly hit Nigeria).

Further, stigmatization can be perceived differently by psychiatric outpatients, and our study did not explore the dimensions of stigmatization that may influence mental health help-seeking behavior. This, therefore, limits the interpretation of our results. Another limitation of the study was the possibility of response bias from participants. Lastly, employing a qualitative study as a follow-up study to explore why religious affiliation significantly predicted mental health help-seeking behavior might have expanded this study (although this was not an objective of our study). These limitations, therefore, mirror multiple hypotheses that can benefit future research.

## CONCLUSION

We concluded that some significant predictors of mental health help-seeking behavior among some psychiatric outpatients attending a Nigerian psychiatric facility have been identified by our research. Therefore, further research that strengthens an interdisciplinary approach to psychiatric care and enhances mental health help-seeking behavior among psychiatric outpatients is needed using a large sample size that is representative.

## Data Availability

Data is available from the corresponding author upon reasonable request.

## ACKNOWLEDGEMENT

To the participants for giving up their time to participate in this study, and to Saada, Gift, and Tayo for their supports on this study.

## INFORMED CONSENT

Both written and verbal consents were used to obtain consent from participants and all participants consented that their data should be used for this research.

## DATA AVAILABILITY STATEMENT

The data that supports this study is available from the corresponding author upon reasonable request.

